# Continued High Mortality Following Diagnosis of HIV-Associated Kaposi Sarcoma in East Africa in the “Treat All” Antiretroviral Therapy Era ― 2021-2024

**DOI:** 10.64898/2026.03.10.26348056

**Authors:** Helen Byakwaga, Aggrey Semeere, Megan Wenger, Esther Freeman, Miriam Laker-Oketta, Elyne Rotich, Beatrice P. Mushi, Matthew Ssemakadde, Hilda Muwando, Bronia Mwine, Racheal Ayanga, Celestine Lagat, Sigrid Collier, Zainab Illonga, Robert Lukande, Philippa Kadama-Makanga, Pendo Ibrahim, Linda Chemutai, Toby Maurer, Charles Kasozi, Winnie Muyindike, Elia Mmbaga, David V. Glidden, Samson Kiprono, Kara Wools-Kaloustian, Andrew Kambugu, Jeffrey Martin

**Affiliations:** Infectious Diseases Institute, Makerere University College of Health Sciences, Kampala, Uganda; University of California, San Francisco; Harvard Medical School, Boston, Massachusetts; Academic Model Providing Access to Healthcare, Eldoret, Kenya; Muhimbili University of Health and Allied Sciences, Dar es Salaam, Tanzania; Ministry of Health, Masaka Regional Referral Hospital, Uganda; Mbarara University of Science and Technology, Uganda; University of Washington, Seattle, Washington; Veterans Affairs Puget Sound Health Care System, Seattle, Washington; Indiana University, Indianapolis, Indiana; Moi University, Eldoret, Kenya

**Keywords:** Kaposi sarcoma, survival, HIV, staging, East Africa, Africa, Kenya, Uganda, Tanzania

## Abstract

**Background:** Stage at time of diagnosis and survival after diagnosis are critical parameters regarding control of any cancer in any geographical setting. In earlier research focusing on the initial years of the “Treat All” era (2016-2019), we found that HIV-associated Kaposi sarcoma (KS) in East Africa continued to be diagnosed at advanced stage of disease and conferred high mortality. Given the potential for broader implementation of “Treat All” as well as the announcement of National Comprehensive Cancer Network guidelines for cancer treatment in Africa since 2020 — but also the countervailing influence of the COVID-19 pandemic — we sought to provide an update on KS stage at diagnosis and survival after KS diagnosis among people living with HIV (PLWH) in East Africa.

**Methods:** We evaluated adult PLWH in Kenya, Tanzania and Uganda with a new diagnosis of KS identified at ambulatory and inpatient settings in four regions between September 2021 and April 2024. At time of biopsy, participants were examined to document the extent of KS. In a prospective cohort study, we followed participants to monitor vital status.

**Results:** Among 493 PLWH with a new diagnosis of KS, the median (IQR) number of anatomic sites with KS lesions was 9 (4-12), and 91% had ACTG stage T1 (advanced KS). Over a median follow-up of 11 (IQR: 2.2-20) months, a total of 209 participants died, and three were lost to follow-up. Cumulative incidence of death (95% confidence interval) at months, 3, 6, 12 and 18 following KS diagnosis was 26% (22% to 30%), 32% (28%-36%), 39% (34%-43%) and 45% (40%-51%), respectively. Cumulative incidence of death was similar between countries and year of KS diagnosis.

**Conclusions:** Among PLWH with newly diagnosed KS in East Africa during the post-initial phase of the “Treat All” era (2021-2024), the majority had advanced disease at KS diagnosis and survival was very poor. These parameters are unchanged from the five prior years. Our findings emphasize the need for better KS control strategies in the region, including primary prevention, novel approaches for earlier detection, more timely linkage to care, and more accessible and potent anti-KS therapy.

## Background

The World Health Organization (WHO)’s “Treat All” strategy seeks to initiate antiretroviral therapy (ART) for all individuals diagnosed with HIV, regardless of clinical stage or CD4 count, with a goal of preventing serious morbidity and mortality.^1^ While this approach, announced in 2015, has been successful in reducing overall HIV-related hospitalizations and mortality in most resource-limited settings,^2,3^ its impact on specific HIV-associated conditions remains less well understood. In earlier research focusing on the initial years of the “Treat All” era (2016 to 2019) in East Africa, we found that one condition — Kaposi sarcoma (KS) — continued to be diagnosed at advanced stage of disease and conferred high mortality.^4^ During this period, among people living with HIV (PLWH) newly diagnosed with KS in Kenya and Uganda, 91% had advanced stage KS at the time of KS diagnosis and 41% died by one year after KS diagnosis.

The five years that have followed our initial report have provided additional time for “Treat All” to potentially become more widely implemented, as well as time for adoption of the National Comprehensive Cancer Network (NCCN) treatment guidelines for KS.^5^ However, this period has also been marked by the COVID-19 pandemic, a major disruption for many aspects of clinical care, including early cancer detection and timely cancer treatment.^6,7^ To date, we are unaware of how these circumstances have collectively influenced the control of KS in East Africa. Therefore, we sought to provide an update on stage and survival following a diagnosis of KS in a community-derived sample of PLWH in East Africa during the period 2021 to 2024.

## Methods

### Overall Design

In a prospective cohort study, we enrolled PLWH who were newly diagnosed with KS at several regions in East Africa. Physical examination at the time of diagnosis provided information on staging and subsequent follow-up estimated survival after diagnosis.

### Study Population

*Study sites.* Programs in four regions participated: a) Academic Model Providing Access to Healthcare (AMPATH) network in western Kenya; b) Masaka Regional Referral Hospital (Masaka-RRH), and the Infectious Diseases Institute (IDI), Kampala, in central Uganda; c) Mbarara Regional Referral Hospital (Mbarara-RRH) in south-western Uganda; and d) Muhimbili National Hospital, Dar es Salaam, in the eastern coastal region of Tanzania. These programs, of which a) through c) contributed to our earlier research, were chosen because they offered free-of-charge skin biopsy services for patients with suspected KS. These services attract referrals not only from existing patients at the ambulatory and inpatient units within the participating facilities but also from individuals originating from the community either out of care or receiving care at facilities that lack biopsy capacity.^4,8^ This context provided what we believe is the best opportunity in East Africa to investigate a community-representative sample of new diagnoses of HIV-associated KS.

*Identification of study participants.* Between September 2021 and April 2024, we approached consecutive adult (≥ 18 years) PLWH, who presented to or were referred to each facility’s respective skin biopsy service with lesions sufficiently suspicious for KS to warrant a biopsy, for their interest in participating in our research. Those who agreed and whose subsequent biopsies revealed KS by local pathologists were enrolled and subsequently followed. Anti-latency-associated nuclear antigen (LANA) immunohistochemical staining was used to confirm diagnosis of KS, at the discretion of the pathologist. For the rare instances where the only lesions present were deemed not safe to biopsy (e.g., conjunctival or oral lesions), a clinical diagnosis of KS was made based on the characteristic macroscopic appearance and agreement of two or more clinicians.

### Measurements

*Questionnaire-based.* At time of biopsy, interviewer-administered questionnaires ascertained socio-demographic characteristics, history regarding HIV infection, and information on symptoms referable to KS, including those suggestive of gastrointestinal or pulmonary KS involvement. During follow-up, information was collected on vital status and chemotherapy use. We intentionally limited measurements during follow-up to minimize research-level intrusion and thereby capture real-world survival.

*Physical examination.* Oral and skin examinations were performed at enrollment to determine the extent of KS by assessing the number of anatomic regions with lesions suspected to be KS, total number of KS-suspicious lesions, macroscopic lesion morphology, and presence of edema.

*Laboratory tests.* At enrollment, we performed complete blood count, plasma HIV RNA viral load (Amplicor HIV Monitor version 1.5 or the Cobas Taqman HIV-1 version 1.0 assays; Roche) and CD4+ T-cell counts (FACSCalibur; Becton Dickinson).

*Participant follow-up.* Following enrollment, participants were evaluated with in-person or phone-administered interviews: first within 2 weeks of the biopsy, then every 3 months in the first year, and every 6 months thereafter. We used previously described methods to perform community tracing^9^ for participants who missed study visits. The study did not administer any KS-specific therapy but instead referred participants to respective oncology units. Chemotherapy was recommended, if available and accessible by patients, for all those with advanced disease, and on case-by-case basis, for those with less than advanced disease. Likewise, the study did not administer HIV therapy; participants were either referred to facility-affiliated HIV care clinics or continued to receive treatment from their care providers.

### Statistical Analysis

The Kaplan-Meier approach was used to estimate survival after KS diagnosis. Time zero was defined as the date of biopsy. defined as duration from time zero to either the date of death or the date last date known to be alive (for those not known to have died) up to the date of administrative censoring (December 30, 2024). Stata 15 (College Station, Texas) was used for analysis.

## Results

### Characteristics of Adults with Newly-Diagnosed HIV-Associated KS

Between September 2021 and April 2024, 693 PLWH were evaluated by the biopsy services at the participating healthcare facilities in East Africa because of mucocutaneous lesions that were clinically suspicious for KS (Figure 1). Of these, 10 patients either declined a biopsy or the procedure could not be immediately performed. Among the 683 PLWH in whom a biopsy was performed, 524 were histopathologically confirmed to have KS. Of these, 31 were not enrolled in the study because of inability to provide consent, declined participation, or the study team lacked time to complete enrollment (for example, due to patient presentation later in the day). Of the 493 patients who consented to participate, 126 (26%) were identified at the AMPATH network in Kenya; 44 (8.9%) at MUHAS in Tanzania; and 250 (51%) at the IDI, 41 (8.3%) at Masaka-RRH, and 32 (6.5%) at Mbarara-RRH in Uganda. Regarding year of enrollment, 5.5%, 44%, 41%, and 9.5% were diagnosed with KS in 2021, 2022, 2023 and 2024, respectively. The majority of participants (66%) were men, the median (interquartile range, IQR) age was 35 (29-42) years, and monthly household income was 53 (14-108) U.S. dollars (Table 1). On physical examination, the median number of anatomic sites with KS lesions was 9 (IQR: 4 to 12), 39% had ≥50 distinct KS-suspicious skin lesions, 23% had ulceration on KS-suspicious skin lesions, 83% had KS-associated edema, 58% had oral KS-suspicious lesions, and 91% had advanced KS (AIDS Clinical Trials Group (ACTG) Tumor 1 (T1) stage).^10^ Upon laboratory testing on the date of diagnosis, 22%, 35%, 34% and 9.0% of participants had CD4+ T cell counts less than 50 cells/µl, 51 to 200 cells/µl, 201 to 500 cells/µl, and greater than 500 cells/µl, respectively.

**Figure 1.**
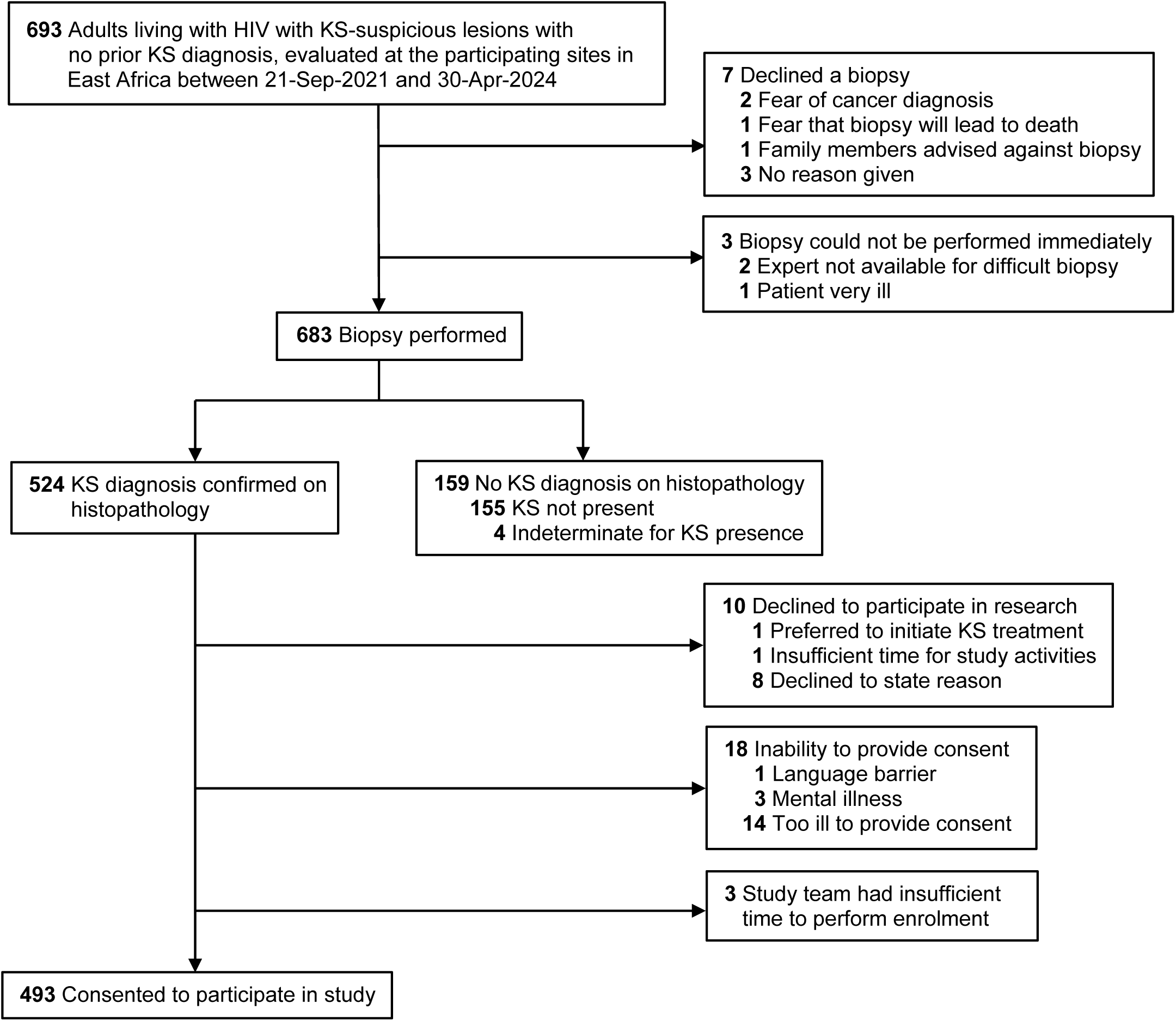
Identification of study participants. This prospective cohort study enrolled adults living with HIV infection who had a biopsy-confirmed new diagnosis of Kaposi sarcoma (KS) recorded between 21 September 2021 and 30 April 2024 at one the five participating sites in Kenya, Tanzania, and Uganda. Two individuals for whom a biopsy could not be performed immediately had KS-suspicious lesions in anatomical locations (e.g., oral cavity or conjunctivae) that were deemed to require a surgical expert for safe biopsy but for whom an expert was not available during the initial encounter and the patient did not return and could not be located for additional attempts.

**Table 1.**
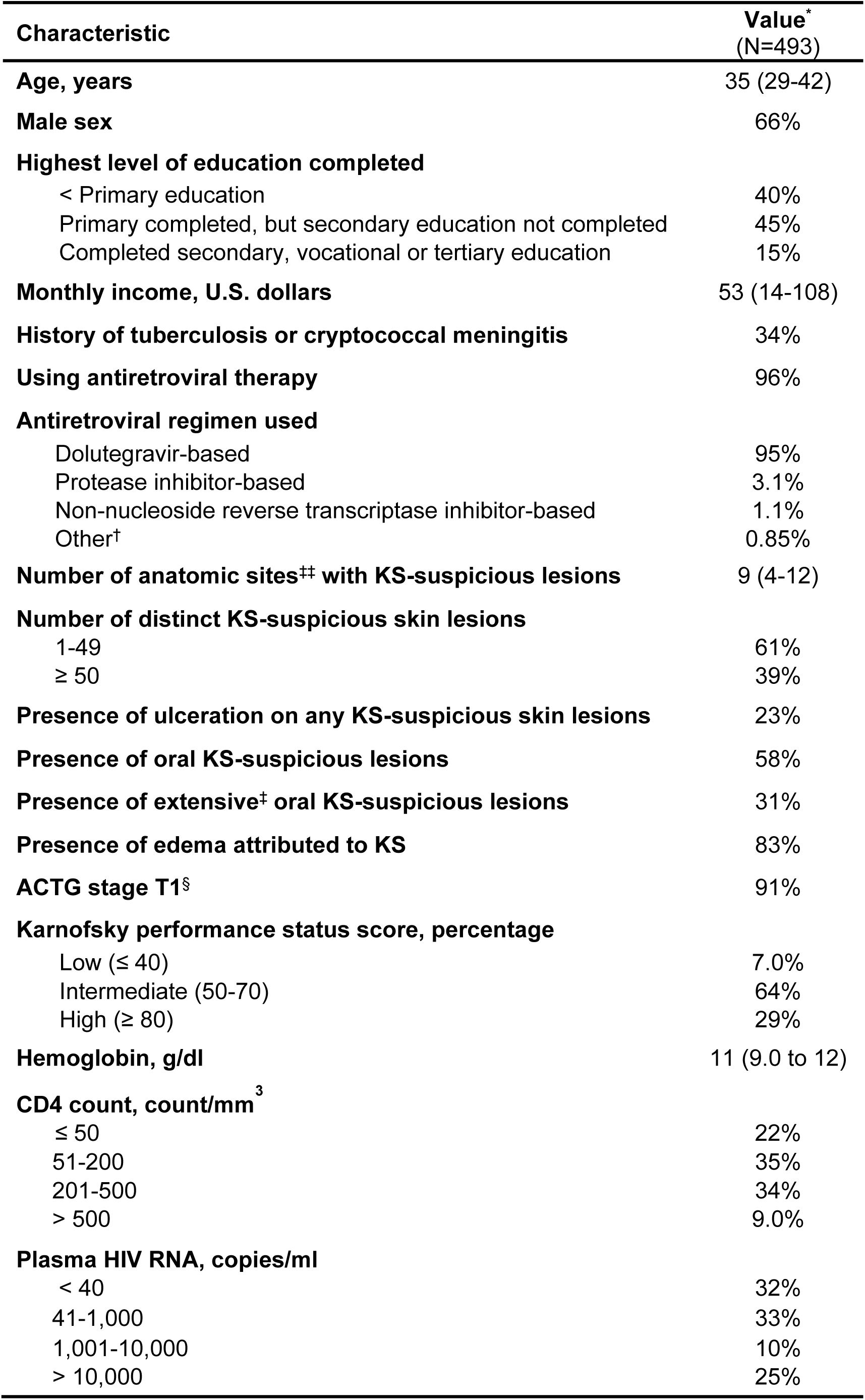
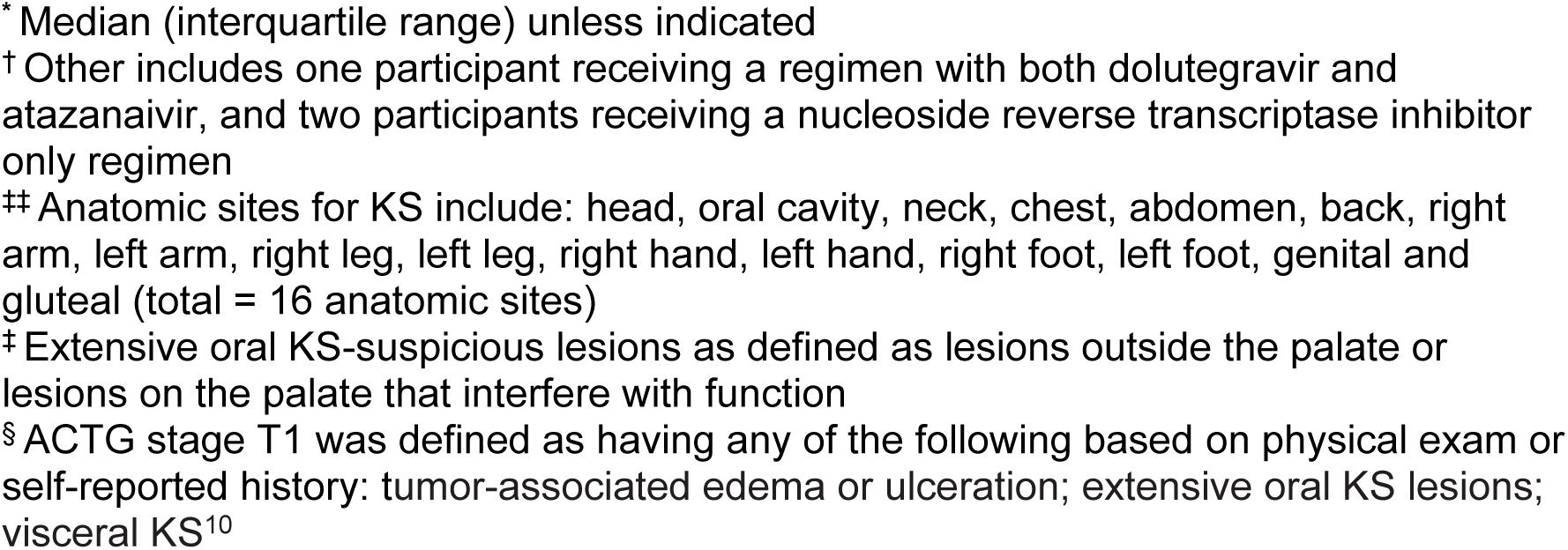
Sociodemographic and clinical characteristics at the time of diagnosis of Kaposi sarcoma (KS) among adults living with HIV who were newly diagnosed with KS in Kenya, Tanzania and Uganda from 2021 to 2024.

| **Characteristic** | **Value\***<br>**(N=493)** |
| --- | --- |
| **Age, years** | 35 (29-42) |
| **Male sex** | 66% |
| **Highest level of education completed** |  |
| < Primary education | 40% |
| Primary completed, but secondary education not completed | 45% |
| Completed secondary, vocational or tertiary education | 15% |
| **Monthly income, U.S. dollars** | 53 (14-108) |
| **History of tuberculosis or cryptococcal meningitis** | 34% |
| **Using antiretroviral therapy** | 96% |
| **Antiretroviral regimen used** |  |
| Dolutegravir-based | 95% |
| Protease inhibitor-based | 3.1% |
| Non-nucleoside reverse transcriptase inhibitor-based | 1.1% |
| Other<sup>†</sup> | 0.85% |
| **Number of anatomic sites<sup>‡‡</sup> with KS-suspicious lesions** | 9 (4-12) |
| **Number of distinct KS-suspicious skin lesions** |  |
| 1-49 | 61% |
| ≥ 50 | 39% |
| **Presence of ulceration on any KS-suspicious skin lesions** | 23% |
| **Presence of oral KS-suspicious lesions** | 58% |
| **Presence of extensive<sup>‡</sup> oral KS-suspicious lesions** | 31% |
| **Presence of edema attributed to KS** | 83% |
| **ACTG stage T1<sup>§</sup>** | 91% |
| **Karnofsky performance status score, percentage** |  |
| Low (≤ 40) | 7.0% |
| Intermediate (50-70) | 64% |
| High (≥ 80) | 29% |
| **Hemoglobin, g/dl** | 11 (9.0 to 12) |
| **CD4 count, count/mm<sup>3</sup>** |  |
| ≤ 50 | 22% |
| 51-200 | 35% |
| 201-500 | 34% |
| > 500 | 9.0% |
| **Plasma HIV RNA, copies/ml** |  |
| < 40 | 32% |
| 41-1,000 | 33% |
| 1,001-10,000 | 10% |
| > 10,000 | 25% |

\* Median (interquartile range) unless indicated
† Other includes one participant receiving a regimen with both dolutegravir and atazanavir, and two participants receiving a nucleoside reverse transcriptase inhibitor only regimen
‡ Anatomic sites for KS include: head, oral cavity, neck, chest, abdomen, back, right arm, left arm, right leg, left leg, right hand, left hand, right foot, left foot, genital and gluteal (total = 16 anatomic sites)
‡ Extensive oral KS-suspicious lesions as defined as lesions outside the palate or lesions on the palate that interfere with function
§ ACTG stage T1 was defined as having any of the following based on physical exam or self-reported history: tumor-associated edema or ulceration; extensive oral KS lesions; visceral KS<sup>10</sup>

### Mortality among Adults with Newly-Diagnosed HIV-Associated KS

Over a median follow-up of 11 months (absolute range: 0.02 to 39; IQR: 2.2 to 20), only 3 participants were lost to follow-up with regard to vital status, i.e., the study could not ascertain their vital status at the time of administrative closure. During follow-up, 55% participants were administered at least one dose of chemotherapy; chemotherapeutic drugs initiated were paclitaxel (56%), liposomal doxorubicin (26%), bleomycin plus vincristine (3%), etoposide or pomalidomide (1%) and drug information was unknown for 13% of patients). A total of 209 deaths occurred, corresponding to a cumulative incidence of death (95% confidence interval (CI)) at months 3, 6, 12 and 18 following KS diagnosis of 26% (22% to 30%), 32% (28% to 36%), 39% (34% to 43%) and 45% (40% to 51%), respectively (Figure 2A). There was no strong evidence of a difference between countries (Figure 2B) or between year of KS diagnosis (Figure 2C).

**Figure 2.**
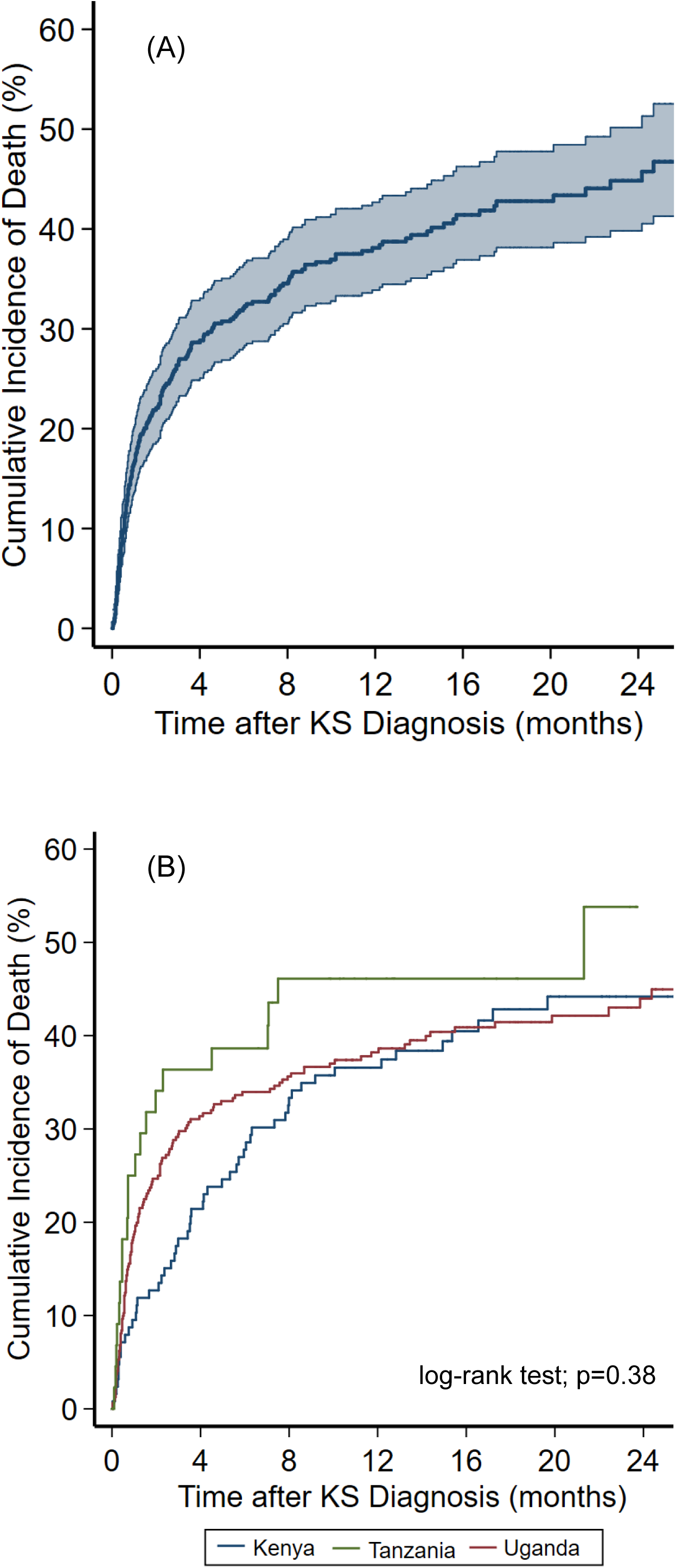
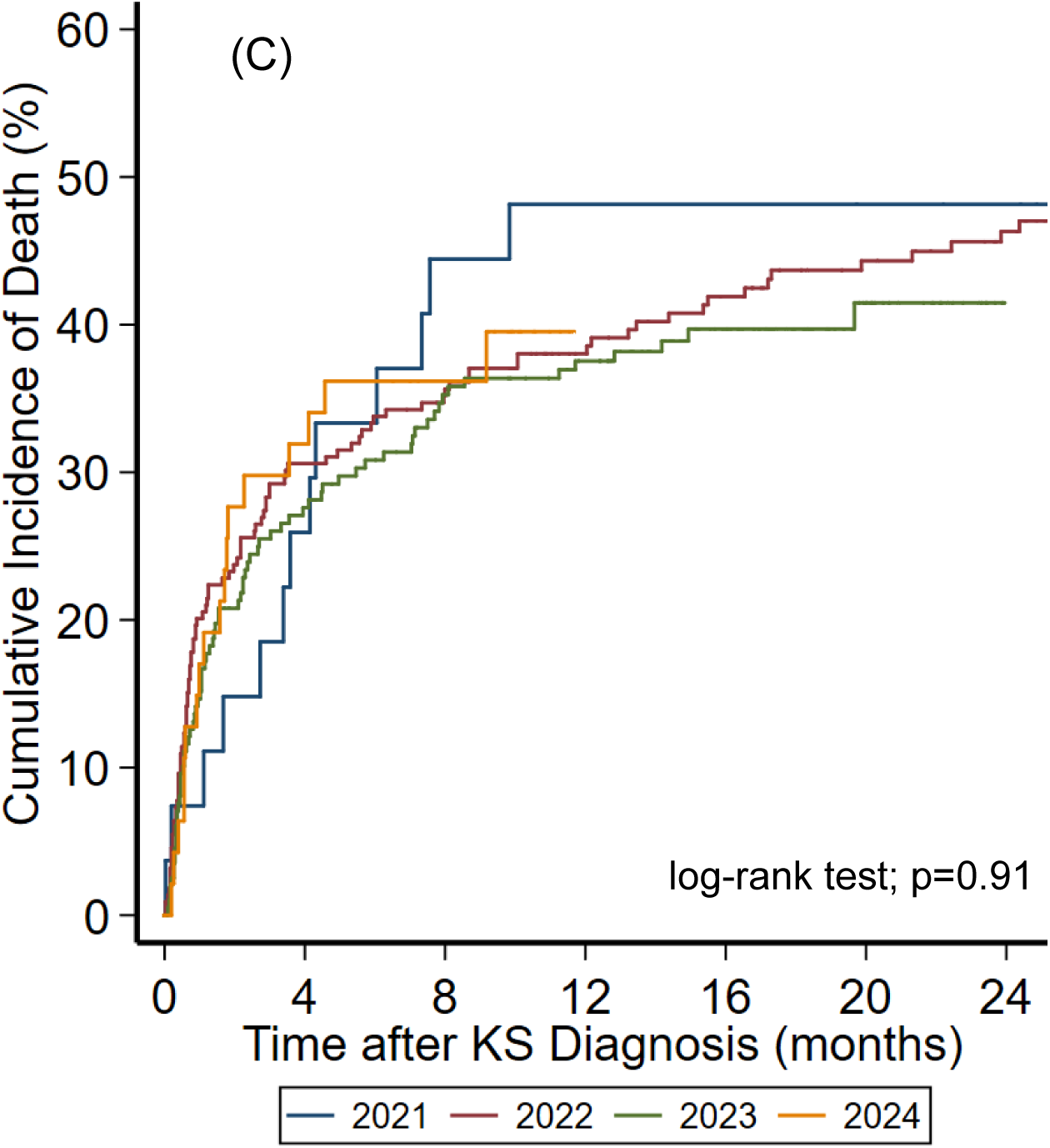
Cumulative incidence of death following diagnosis of KS among adults living with HIV with newly diagnosed Kaposi sarcoma between 2021 and 2024 at participating health facilities in East Africa. All estimates were determined by the Kaplan-Meier estimator in which participants who became lost to follow-up were censored on the last date they were last known to be alive. Panel A depicts the estimation for the entire study population. The shaded area represents the 95% confidence interval around the cumulative incidence estimate. Panel B depicts cumulative incidence of death by participant’s country of residence: Kenya, Tanzania, and Uganda. Panel C depicts cumulative incidence of death by the year of KS diagnosis: 2021 to 2024.

## Discussion

Stage at diagnosis and survival after diagnosis are critical parameters regarding the control of cancer in any setting. Stage integrates patient willingness/ability to access care and the health care system’s ability to recognize and diagnose cancer (particularly in its earlier stages), while survival captures both biologic aggressiveness of the cancer and the health care system’s ability to deliver effective treatment. Our prior reports from the early years of the “Treat All” era demonstrated a remarkably high frequency of advanced disease at time of HIV-related KS diagnosis in East Africa as well as poor survival.^4,11^ These findings urged continued monitoring. Among PLWH in East Africa diagnosed with KS between 2021–2024, we found most patients continue to have advanced-stage disease at time of KS diagnosis, and survival remains poor. These findings are unchanged compared to the five preceding years.

In contrast to resource-rich settings with publicly-funded cancer registries that tally all incident cancer diagnoses (and cancer stage at time of diagnosis) and death registries that comprehensively tally deaths ― thus allowing for accurate monitoring of cancer survival — most resource-limited countries lack the infrastructure necessary to generate comparable data on stage and survival after a cancer diagnosis. As a result, we are not aware of any data during the period of our work regarding KS stage at diagnosis or survival in sub-Saharan Africa to which we can compare our results. We expect that the nominally population-based cancer registries in Africa will eventually report updated estimates of KS survival, but, in the past, losses to follow-up have threatened validity,^12^ a threat we avoided by expending considerable effort on community tracing.^9^ Thus, in the near future, it is likely that only independent research-dedicated efforts, similar to what we have performed, will be able to derive stage and survival estimates for KS to which our findings can be compared.

There are several potential explanations for why survival after KS diagnosis remains poor. First, the majority of PLWH continue to be diagnosed with advanced KS, which has been consistently associated with poor survival.^4,13,14^ The reasons for late stage at time of diagnosis include both patient delay and health system failure for diagnosis.^15^ Second, although chemotherapy improves survival after KS diagnosis, substantial barriers to accessing oncologic care remain,^16^ and a large fraction of patients for whom chemotherapy is indicated never initiate treatment.^17^ Third, persistently poor survival after Kaposi sarcoma diagnosis may reflect the limited effectiveness of available therapies. Compared to our prior work,^4^ there was a shift towards use of paclitaxel and liposomal doxorubicin instead of bleomycin plus vincristine, aligning with current NCCN guideline recommendations for KS management in sub-Saharan Africa.^5^ However, survival following KS diagnosis remained unchanged, despite the use of these agents with nominally better efficacy and tolerability. This is consistent with trial data showing substantial mortality despite optimal therapy, underscoring that current treatments are not curative.^18^ The attributable risk of these different explanations for poor survival (i.e., which is most important) is not known, and until a time when this is known, it seems prudent to consider more research and solutions for all of the deficiencies. Each is a difficult problem, but we believe that primary and secondary prevention may be more tractable than finding better more potent therapies.

Another potential explanation for poor survival in this patient population is because they harbor other concurrent fatal diagnoses (e.g., opportunistic infections or other malignancies). Indeed, the ideal metric to assess the impact of HIV-associated KS on survival would compare PLWH with KS to a counterfactual group of HIV-infected individuals without KS (i.e., the same people but without KS).

In practice, of course, such a counterfactual cannot be directly observed. The best approximation is to identify incident cases of HIV-associated KS and compare them to HIV-infected persons without KS who have similar serious concurrent conditions. In resource-limited settings, however, diagnostic capacity is often insufficient, and many comorbid conditions go undetected. This unmeasured concurrent morbidity is the source of residual unmeasured confounding, which is essentially impossible to mitigate. An alternative design is to select a non-KS comparator group of HIV-infected persons similar in CD4+ T cell count and plasma HIV RNA level and presumed to have no other conditions (e.g., not hospitalized and no symptoms). Although this group can be identified with reasonable confidence, it is not the correct comparator because it does not reflect the true burden of concurrent illness present in patients with KS. Comparisons to HIV-infected groups with other confirmed opportunistic infections (e.g., tuberculosis or cryptococcosis), adjusting for CD4 and HIV RNA, can provide a lower-bound estimate of KS-attributable mortality; however, this is imperfect because the overall KS group is not limited to patients with at least one other serious condition. With the difficulties posed by all these approaches, we concede that valid estimation of the fraction of death strictly attributable to KS among patients with newly diagnosed HIV-related KS is very hard in resource-limited settings.

While survival from HIV-associated KS in East Africa has been unchanged over the past 9 years, survival alone tells an incomplete depiction. Total KS burden is a product of both survival and incidence.^19^ Truth regarding incidence of HIV-associated KS in the region, however, has been elusive because accurate capture of all KS that arises from a representative enumerated population of PLWH is difficult. A recent review summarized available KS incidence data from nominal population-based sources.^20^ It estimated a 27% decline (but without statistical significance) in KS incidence between 2001–2010 and 2011–2016. This review acknowledged significant under-reporting of cancer incidence, and others have also criticized estimates in resource-limited regions.^21,22^ Consequently, there remains uncertainty about the true magnitude of KS decline in Africa. Although exact magnitude of decline is unknown, what is true is that it does not appear to approach the nearly 90% diminution seen in resource-rich settings,^23–25^ and KS remains, in the most recent data from 2022, among the most commonly diagnosed cancers in the general population in several countries in Eastern Africa.^26^ In short, the faucet of KS incidence in sub-Saharan Africa is far from closed.

There are limitations to our work. In our prior work, we scoured participating sites for all KS diagnoses by regularly searching histopathology logs and medical records of inpatient wards and HIV primary care, dermatology and oncology clinics, in addition to monitoring each facility’s skin biopsy service.^4,8^ In contrast, in the current study, we only approached patients who interfaced with the biopsy services; these were persons referred from the participating facility’s inpatient and ambulatory units, as well as originating from the community from all of the many facilities without biopsy capacity. The longstanding presence of these biopsy services and referrals made to them from other units provided confidence that the current approach did not miss diagnoses, but we lack direct proof that no diagnoses were missed. Yet, if diagnoses were missed, it is not obvious that they would be enriched for either early or advanced disease. In addition, as in our earlier study, we possibly missed some KS that arose from the community and was never diagnosed either because individuals never presented for care and died at home or presented to care and died due to comorbid conditions before KS diagnosis was made. For these reasons, while we believe our community-derived sample is as close to population-representative as realistically feasible, it may not be pristinely representative.

In summary, among PLWH in East Africa with newly diagnosed KS between 2021 to 2024, we found that despite being nearly 10 years removed from the start of “Treat All”, the majority had advanced disease at the time of KS diagnosis and had poor survival. Our findings re-emphasize the need for better KS control strategies, including primary prevention, earlier detection, more timely linkage to care, and more accessible and potent anti-KS therapy.

## List of Abbreviations

ACTG: AIDS Clinical Trials Group
AMPATH: Academic Model Providing Access to Healthcare
ART: Antiretroviral therapy
CI: Confidence interval
IDI: Infectious Diseases Institute
IQR: Interquartile range
KS: Kaposi sarcoma
LANA: Latency-Associated Nuclear Antigen
Masaka-RRH: Masaka Regional Referral Hospital
Mbarara-RRH: Mbarara Regional Referral Hospital
MUHAS: Muhimbili University of Health and Allied Sciences
NCCN: National Comprehensive Cancer Network
PLWH: People living with HIV
REC: Research Ethics Committee
T1: Tumor 1

## Declarations Ethics approval and consent to participate

This study received ethics approval in Kenya: from Moi University Research Ethics Committee (REC) ─ (Reference: IREC/2020/237); in Tanzania, from the National Institute for Medical Research (Reference: NIMR/HQ/R.8a/Vol. IX/4039); and in Uganda, from the Makerere University College of Health Sciences School of Biomedical Sciences Higher Degrees REC (Reference: SBS-HDREC-495). Written informed consent was obtained from all participants.

## Consent for publication

Not applicable

## Availability of data and materials

The datasets used are available from the corresponding author on reasonable request.

## Authors’ contribution

HB: Conceptualization, Methodology, Investigation, Formal analysis, Writing-Original Draft, Project administration, Visualisation, Supervision. AS: Conceptualization, Methodology, Investigation, Writing-Review and Editing, Project administration, Supervision. MW: Software, Data Curation, Writing-Review and Editing. EF: Methodology, Writing-Review and Editing. MLO: Methodology, Writing-Review and Editing, Project administration. ER: Project administration, Data curation, Writing-Review and Editing. BPM: Project administration, Writing-Review and Editing. MS: Project administration, Data Curation, Writing-Review and Editing. HM: Investigation, Project administration, Writing-Review and Editing. BM: Project administration, Data Curation, Writing-Review and Editing. RA: Investigation, Project administration, Writing-Review and Editing. CL: Investigation, Writing-Review and Editing. SC: Methodology, Writing-Review and Editing. ZI: Investigation, Project administration, Writing-Review and Editing. RL: Investigation, Validation, Writing-Review and Editing. PKM: Project administration, Writing-Review and Editing. PI: Investigation, Project administration, Writing-Review and Editing. LC: Investigation, Writing-Review and Editing. TM: Methodology, Writing-Review and Editing. CK: Supervision, Writing-Review and Editing. WM: Supervision, Writing-Review and Editing. EM: Supervision, Writing-Review and Editing. DVG: Methodology, Writing-Review and Editing. KWK: Funding, Writing-Review and Editing. SK: Validation, Supervision, Writing-Review and Editing. AK: Funding, Supervision, Writing-Review and Editing. JM: Funding, Conceptualization, Methodology, Writing-Review and Editing, Funding acquisition, Validation, Supervision.

## Data Availability

All data produced in the present study are available upon reasonable request to the authors

## Acknowledgements

We thank all the research staff who contributed to data collection at the different facilities that participated in this study. AMPATH: Philip Odhiambo, Fred Shiravika, and Emily Mulanda; IDI: Jane Frances Nalubega, Prossy Nannyange, and Andrew Mulooki; Masaka-RRH: Haruna Semuwemba and Stella Nabunya; Mbararra-RRH: Martin Mwebesa and Placidia Oinembabazi. MUHAS: Selekwa Msiba.

## Previous presentation

This work was previously presented in part at the 30th Conference on Retroviruses and Opportunistic Infections (CROI), held February 19-22, 2023, in Seattle, Washington, and at the 19th International Conference on Malignancies in AIDS and Other Acquired Immunodeficiencies in Bethesda, Maryland, on October 24-25, 2024.

